# Comparing microbiological and molecular diagnostic tools for the surveillance of anthrax

**DOI:** 10.1101/2024.04.02.24305203

**Authors:** Sunday O. Ochai, Ayesha Hassim, Edgar H. Dekker, Thuto Magome, Kgaugelo E. Lekota, S. Marcus Makgabo, Lin-Mari de Klerk-Loris, O. Louis van Schalkwyk, Pauline L. Kamath, Wendy C. Turner, Henriette van Heerden

## Abstract

The diagnosis of anthrax, a zoonotic disease caused by *Bacillus anthracis* can be complicated by detection of closely related species. Conventional diagnosis of anthrax involves microscopy, culture identification of bacterial colonies and molecular detection. Genetic markers used are often virulence gene targets such as *Bacillus anthracis* protective antigen (*pagA,* as also called BAPA, occurring on plasmid pXO1), lethal factor (*lef,* on pXO1), as well as chromosomal (Ba-1) and plasmid (capsule-encoding *cap*B/C, located on pXO2). Combinations of genetic markers using real-time/quantitative polymerase chain reaction (qPCR) are used to confirm *B. anthracis* from culture but can also be used directly on diagnostic samples to avoid propagation and its associated biorisks and for faster identification. We investigated how the presence of closely related species could complicate anthrax diagnoses with and without culture to standardise the use of genetic markers using qPCR for accurate anthrax diagnosis. Using blood smears from 2012-2020 from wildlife mortalities (n=1708) in Kruger National Park in South Africa where anthrax is endemic, we contrasted anthrax diagnostic results based on qPCR, microscopy, and culture. From smears, 113/1708 grew bacteria in culture, from which 506 isolates were obtained. Of these isolates, only 24.7% (125 isolates) were positive for *B. anthracis* based on genetic markers or microscopy. However, among these, merely 4/125 (3.2%) were confirmed *B. anthracis* isolates (based on morphology, microscopy, and sensitivity testing to penicillin and gamma-phage) from the blood smear, likely due to poor survival of spores on stored smears. This study identified *B. cereus sensu lato*, which included *B. cereus* and *B. anthracis, Peribacillus spp*., and *Priestia spp.* clusters using *gyr*B gene in selected bacterial isolates positive for BAPA. Using qPCR on blood smears, 52.1% (890 samples) tested positive for *B. anthracis* based on one or a combination of genetic markers which included the 25 positive controls. Notably, the standard *lef* primer set displayed the lowest specificity and accuracy. Interestingly, various marker combinations, such as Ba-1+*capB*, BAPA+*capB*, Ba-1+BAPA+*capB*+*lef*, and BAPA+*lef*+*capB*, all demonstrated 100.0% specificity and 98.7% accuracy, while maintaining a sensitivity of 96.6%. The BAPA+*lef*+Ba-1 combination showed 100% specificity, sensitivity, and accuracy. Using Ba-1+BAPA+*lef*+*capB*, as well as Ba-1+BAPA+*lef* with molecular diagnosis accurately detects *B. anthracis* in the absence of bacterial culture. Systematically combining microscopy and molecular markers holds promise for notably reducing false positives, thereby significantly enhancing the detection and surveillance of diseases like anthrax in southern Africa and beyond and reducing the need for propagation of the bacteria in culture.

## Introduction

Anthrax is an ancient zoonotic disease with a documented history dating back to the Bible (Ben-Noun, 2002). While the disease affects many host species, herbivorous mammals are most susceptible, with fatalities often observed in wildlife and livestock. In addition, humans are susceptible to anthrax infections, and cases occur largely due to the handling or consumption of carcasses, infected meat, and hides (Kamal et al., 2011, W.H.O., 2008). Anthrax is generally known to be caused by *Bacillus anthracis,* which is an aerobic or facultative anaerobic, non-motile, Gram-positive, rod-shaped bacterium that produces endospores. This bacterium occurs in two forms, the spore form and the vegetative form (Vilas-Bôas et al., 2007). The virulence factors of *B. anthracis* are encoded on two plasmids: pXO1, which is largely responsible for the production of the toxins, and pXO2, which synthesizes the poly-ɣ-D-glutamic acid capsule (Makino et al., 1989, Okinaka, 1999a). The pXO1 plasmid contains genes responsible for the production of protective antigen (PA, often indicated as BAPA), lethal factor (LF) and oedema factor (EF) proteins. These proteins are grouped as AB-toxins. The A form, which consists of the EF or LF, bears the enzymatic activity (Moayeri and Leppla, 2004, Moayeri and Leppla, 2009, Leppla, 1982). The B component consists of PA, which is the receptor-binding component of the lethal toxin (LT) and oedema toxin (ET), and the courier of LF and EF respectively, into the host cells (Smith et al., 1955, Barth et al., 2004, Moayeri and Leppla, 2009, Leppla, 1982). The PA combines with LF to produce LT, which is responsible for the death of macrophages; the PA also combines with the EF to produce the ET that deactivates the phagocytes of the host (Leppla, 1982).

For a century, the laboratory identification of the disease and its causative agent (*B. anthracis*) was believed to be well described and involved microbiological culture, microscopy, and biochemistry. However, in recent decades, new hypotheses with regards to the disease’s presentation, prevention and infective organisms have been elucidated on the African continent (Antonation et al., 2016, Norris et al., 2021, Tamborrini et al., 2011, Klee et al., 2006). There have been reports of a close relationship of pathogenic and non-pathogenic *Bacillus* spp. resulting in serological cross-reactivity (Marston et al., 2016, Zimmermann et al., 2017) which was also reported in the anthrax endemic region in northern Kruger National Park (KNP) (Steenkamp et al., 2018, Ochai et al., 2022). Anti-PA and LT-neutralising antibodies were also detected at higher rates than expected in animals from a low-incidence area of KNP (southern region) (Ochai et al., 2022). Thus we hypothesised that the animals could be reacting to “anthrax-like” microbes in the environment, which could possess genes with similar homologies to *B. anthracis* (Ochai et al., 2022). These data coupled with the discovery of anthrax cases caused by *B. cereus* biovar *anthracis* (Bcbva) strains in west and central Africa (Hoffmann et al., 2017), prompted us to re-assess the robustness of diagnostic tools currently employed in the surveillance for anthrax in southern Africa.

Different methods have been employed in the diagnosis of bacterial zoonoses (such as *B. anthracis*) over the years. These methods include the identification of bacterial culture isolates, microscopic examination of blood smears, molecular diagnosis targeting pathogen genetic markers and serological identification employing antibodies targeting antigens produced by the pathogen. The success of these techniques, however, depends largely on the specificity and sensitivity of the test being employed (Trevethan, 2017). *Bacillus anthracis* is in the phylum Firmicutes, family Bacillaceae, and belongs to the group referred to as the *B. cereus* group. The *B. cereus* group consists of 8 *Bacillus* species (*B. anthracis*, *B. cereus*, *B. thuringiensis*, *B. mycoides*, *B. pseudomycoides*, *B. weihenstephanensis*, *B. cytotoxicus*, and *B. toyonensis)* that have closely related phylogenies (Radnedge et al., 2003, Rasko et al., 2005) as reflected by high similarities in 16S rRNA gene sequences (Ash et al., 1991, Somerville and Jones, 1972) and other genetic markers such as *gyrB* within the *B. cereus* group (Liu et al., 2021). These species also differ in their aetiology, pathogenesis, clinical manifestations and host preferences (Drobniewski, 1993, Rasko et al., 2005, Pilo and Frey, 2011, Helgason et al., 2000, Ehling-Schulz et al., 2019). The *gyrB* gene encodes the B subunit of DNA gyrase, an enzyme critical for DNA replication, transcription, and repair in bacteria. The *gyrB* gene sequence is highly conserved among bacterial species but varies enough to distinguish between them (Yamamoto and Harayama, 1995, Liu et al., 2021). Studies have shown that *gyrB* sequencing can offer higher resolution than the more commonly used 16S rRNA gene sequencing in differentiating closely related bacterial species (La Duc et al., 2004, Fox et al., 1992).

The initial step in the confirmation of *B. anthracis* in an anthrax-suspected carcass is the examination of blood smears stained with either Gram or Giemsa stain to view the rod-shaped bacterium (W.H.O., 2008). The presence of encapsulated square-ended rod-shaped bacteria that react to the polychrome methylene blue stain indicates the presence of *B. anthracis* and warrants a sample to be sent to a reference laboratory for confirmation (W.H.O., 2008). To confirm the presence of *B. anthracis* in the reference laboratories, the samples are cultured on blood agar to check for colony morphology (W.H.O., 2008), and the absence of haemolysis and sensitivity to penicillin and bacteriophages (Turnbull et al., 1998, Turnbull, 1999). For additional verification, real-time/quantitative PCR (qPCR) is conducted for the presence of *pagA/*BAPA, *lef* (W.H.O., 2008), *capB* (pXO2 (W.H.O., 2008)) and/or *Ba-*1 (Zincke et al., 2020) genes that encode for virulence factors including the PA and capsule as well as *B. anthracis* chromosome, respectively. The qPCR targeting *pagA*/BAPA (pXO1) (W.H.O., 2008) and *cap*C (pXO2) regions (W.H.O., 2008) used by Lekota et al. (2016) reported that presence of *cap*C to be inadequate for distinguishing closely related *Bacillus* species from anthrax outbreaks, while Zincke et al. (2020) used *capB, lef* and Ba-1 targets to differentiate *B. anthracis* from *B. cereus sensu stricto.* Although the Ba-1 marker seems distinctive to *B. anthracis*, its validation has been limited to *B. anthracis* strains of *B. cereus sensu stricto*, similar to the case of *lef* (Zincke et al., 2020, Blackburn et al., 2010). Typically, molecular targets involve the use of specific chromosomal regions unique to *B. anthracis* together with virulence factors situated on either pXO1 or pXO2 plasmids serving as markers (W.H.O., 2008, Lekota et al., 2016, Blackburn et al., 2014), due to the genome similarity amongst closely related *Bacillus* spp. (Léonard et al., 1997, Lechner et al., 1998) and *B. anthracis* virulence plasmids or their parts detected in closely related species (Baldwin, 2020, Klee et al., 2006).

In a typical *B. anthracis* investigation, the presence of *B. cereus* group species that are not *B. anthracis* have been viewed as contaminants, and the absence of the genes linked to both pXO1 and pXO2 further leads the investigator to view the closely related species as not of importance (Logan and Turnbull, 1999). As a result, microbes that lack the *B. anthracis*-specific chromosomal gene (Ba-1) or the pXO1 and pXO2 plasmids can be readily overlooked. In recent years, there have been reports of atypical *B. cereus* strains that are known to cause anthrax-like infections in both humans and animals (Klee et al., 2010) with very similar genes to those found on pXO1 and pXO2 plasmids found in *B. anthracis* (Baldwin, 2020). A number of anthrax-like pneumonia cases by atypical *B. cereus* (with the pBCXO1 plasmid) have been reported in humans (Hoffmaster et al., 2004, Avashia et al., 2007, Hoffmaster et al., 2006) with all cases found in individuals without underlying predisposing conditions. Also, cutaneous *B. cereus* infections characterised by the presence of anthrax-like eschars have been reported in humans (Marston et al., 2016, Kaiser, 2011). Furthermore, anthrax-like illnesses have been reported in animals, some of which include chimpanzees (*Pan troglodytes*), gorilla (*Gorilla gorilla*) (Leendertz et al., 2006, Leendertz et al., 2004, Klee et al., 2006, Hoffmann et al., 2017), cattle (*Bos taurus*) (Pilo et al., 2011, Somerville and Jones, 1972), elephants (*Loxodonta africana*) and goats (*Capra hircus*) (Antonation et al., 2016) in west and central Africa. Most of these mortalities have been attributed to atypical strains of *B. cereus* and *B. cereus* biovar *anthracis* (Bcbva). One of the most common diagnostic markers used in the detection of Bcbva is the genomic island IV (GI4) which is unique to Bcbva (Zincke et al., 2020). The review by Baldwin (2020) indicated two *B. cereus* clades that cause anthrax-like disease namely the atypical *B. cereus* strains with pXO1-like-pBCX01 causing human cases of anthrax-like pneumonia (which include G9241, FL2013 and 03BB102), and Bcbva strains containing the pBCXO1 and pBCXO2 plasmids that are highly similar to the pXO1 and pXO2 plasmids of *B. anthracis* (Baldwin, 2020, Okinaka, 1999b, Rasko et al., 2005). Even though atypical *B. cereus* strains possess the ability to cause anthrax-like disease, they are more closely phylogenetically related to other *B. cereus* strains than *B. anthracis* (Antonation et al., 2016). Over the last decades, there have been calls to move away from culture identification of *B. anthracis* in a bid to reduce biosafety risk and avoid proliferation (Riedel, 2005). Thus, the ultimate goal of this study was to investigate the best practices using culture-free methods for the diagnosis of anthrax.

Anthrax is endemic in KNP, and park personnel employ a passive surveillance system where blood smears are collected from any deceased animal and stored in an archival collection. We utilised blood smears from the collection, covering the years 2012-2020. This period encompassed known anthrax outbreaks from 2012 to 2015 (Hassim, 2017a, Ochai et al., 2022). From these outbreaks, *B. anthracis* bacilli were initially identified using the microscopic evaluation of blood smears from wildlife carcasses in KNP with follow-up collection of samples (bone, hair and tissue) from these positive carcass sites in a previous study (Hassim, 2017b). In this study, 25 *B. anthracis* isolates previously confirmed using microbiology and PCR from tissue samples linked to positive blood smears served as positive controls. Our investigation focused on employing microscopy, culture, and molecular markers, including real-time/quantitative polymerase chain reaction (qPCR), to identify *B. anthracis* and distinguish it from *B. cereus* or other closely related microbes. Specifically, we examined: 1) the performance of five molecular markers currently in use (*pagA*/BAPA, Ba-1, *lef, cap*B, *GI*4) to identify *B. anthracis* from other bacteria using cultures of blood smears; (2) the performance of five molecular markers to identify *B. anthracis* from *B. cereus* and other closely related bacteria; and 3) we evaluated the agreement between anthrax diagnoses based on blood smear microscopy versus molecular techniques.

## Materials and Methods

### Study Area

The KNP (19,485 km^2^; Figure 1) is situated in the northeastern part of South Africa, bordering Mozambique and Zimbabwe. The northern half of KNP (Figure 1) is considered the anthrax endemic region, where most of the anthrax mortalities have been reported (Ochai et al., 2022, Vos, 1990); this region is classified as semi-arid and is highly wooded with some grassland savannah (Huntley, 1982). KNP has variable elevations, with Pafuri (found in the northernmost part of KNP; 22.4206° S, 31.2296° E) having lower elevation floodplains and mountains towards the northwestern part of the park. In KNP, the high anthrax incidence (endemic) area extends from Pafuri to Shingwedzi (23.1167° S, 31.4333° E) in the north, and the low incidence area extends from Skukuza (24.9948° S, 31.5969° E) to Crocodile Bridge (25.3584° S, 31.8935° E) in the south (Figure 1).

**Figure 1:**
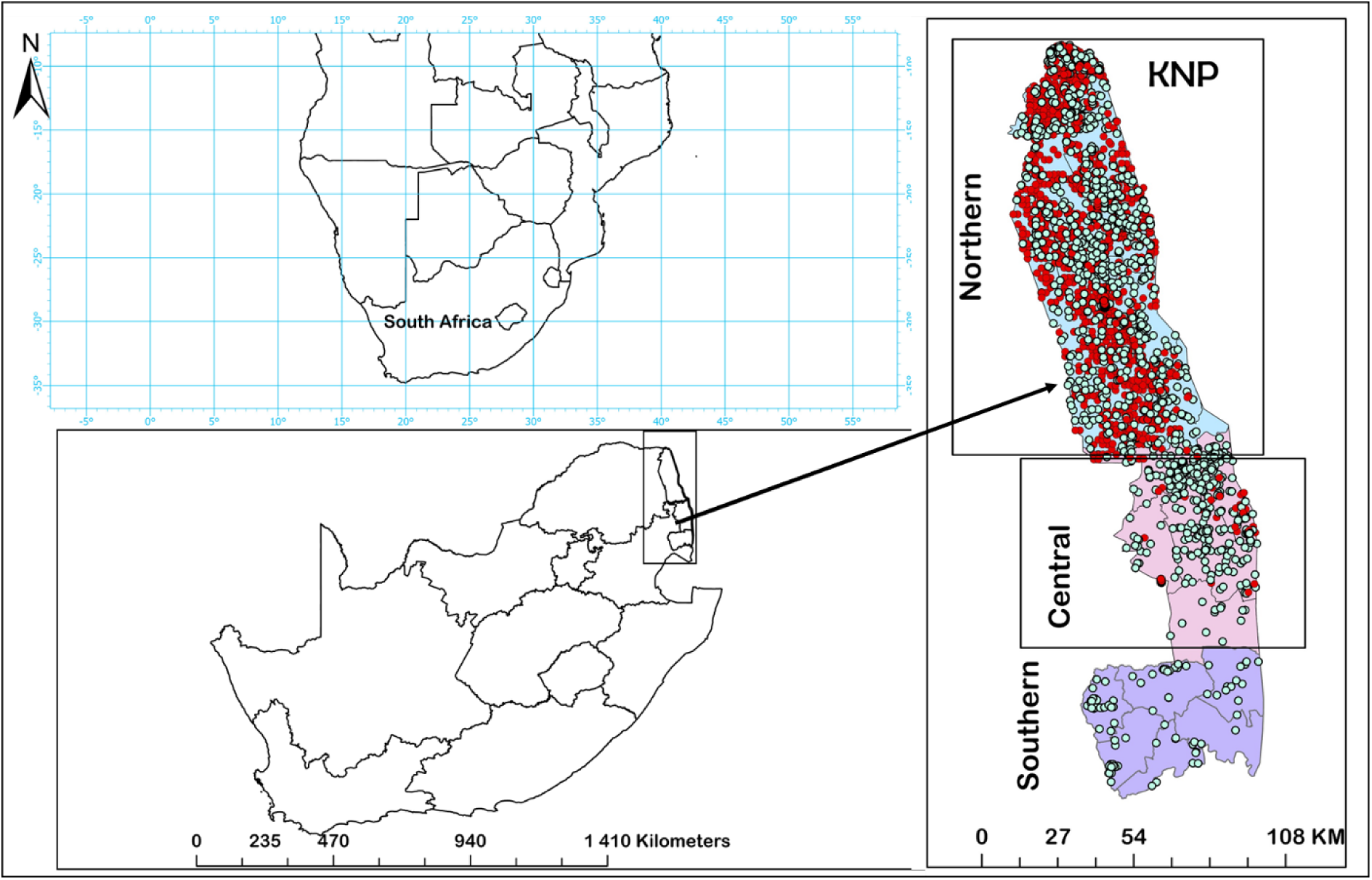
The study area, Kruger National Park (KNP), is located in South Africa. The map of KNP divides the park into three regions, with the distribution of 1708 animal mortalities (Table S1) investigated in this study are shown as dots; anthrax positive cases, identified through microscopic examination of blood smears, are marked with red dots, while green dots indicate anthrax-negative mortalities.

### Sample preparation and DNA extraction

Archival blood smears can be an important resource for retrospective studies and for retrieving pathogens like *B. anthracis* that can remain viable for years (Vince et al., 1998, Hassim, 2017a). In KNP, as part of the passive surveillance by the Skukuza State Veterinary Services, blood smears have been collected from all carcasses discovered during field surveys. Two smears were collected per carcass, one of which is stained (with Giemsa), while the other remains unstained. Metadata captured at the carcass sites include the date, Global Positioning System (GPS) coordinates, locality, species, and sex. These smears were first examined at the time of collection and then stored at room temperature since collection. Aminu et al. (2020) demonstrated that Azure B staining is more robust, consistent and has a higher sensitivity compared to Giemsa only without Azure B and Polychrome Methylene Blue (PMB) stains. The Giemsa stain used in this study contained Azure B.

A total of 1708 Giemsa-stained blood smear slides (from wildlife mortalities recorded 2012-2020; Table S1) were examined by microscopy at 1000X magnification for the presence of square-ended cells indicative of *B. anthracis*, following the methods described by the World Health Organization (W.H.O., 2008). Each slide was examined by two examiners. The selection of smears from this time period (2012-2020) was based on the findings of Hassim (2017a) who demonstrated that isolate recovery reduced with age of the smears. We used the selected unstained smears for additional genetic and microbiological work, done in 2022.

The unstained blood smears from each mortality were scraped into a collection plate and transferred into a 1.5 mL centrifuge tube using a sterile scalpel. The smear scrapings were added to 200 µL of phosphate buffered saline (PBS; Thermo Scientific, MA, USA) and divided into two aliquots. The first aliquot was subjected to automated DNA extraction (QIAcube, QIAGEN GmbH, Hilden, Germany) using the DNA Blood Mini kit (QIAGEN QIAmp, QIAGEN GmbH, Hilden, Germany) and the manufacturer’s instructions for DNA extraction from blood were followed. The second aliquot was inoculated on 5% Sheep Blood Agar (SBA) and incubated overnight at 37 °C for use in the morphological identification of bacterial colonies, as described by Parry et al. (1983). On each plate, all bacterial colonies demonstrating different colony morphology were selected and treated as different isolates. All isolates identified were further sub-cultured onto 5% SBA to obtain pure cultures and check for the presence of haemolysis and colony morphology. The purified isolates were further subjected to gamma-phage and penicillin sensitivity tests as described by W.H.O. (2008). Isolates that did not present with a *B. anthracis* characteristic phenotype were retained and screened using molecular methods. DNA extraction from pure isolates was performed using the Pure link Genomic DNA kit (Thermo Fisher Scientific, MA USA) as prescribed by the manufacturer.

If a mortality was identified as positive for *B. anthracis* based on microscopy, a follow-up sample (soil, bone and/or tissue) from the carcass site was collected as soon as possible (if GPS coordinates were available). From these additional samples, 25 isolates confirmed to be *B. anthracis* based on morphology, microscopy, haemolysis, gamma-phage and penicillin sensitivity (W.H.O., 2008) were used in this study as internal positive controls.

### Microscopic examination of bacterial isolates derived from blood smears

For the blood smear scrapings that yielded bacterial growth, the colonies were subcultured (to obtain pure colonies) and transferred directly to a microscope slide, and 5 μl of saline was added, emulsified, and spread evenly on the slide. The slide was allowed to dry and fixed with 95% methanol (Merck KGaA, Darmstadt, Germany) for one minute. The methanol was allowed to dry and a Gram stain was conducted as described by W.H.O. (2008) to visualise the presence of Gram positive rods. Cell morphology was observed and recorded at 1000X magnification to confirm the culture results to identify square-ended rod-shaped *B. anthracis*. Subsequently, to determine encapsulation, polychrome methylene blue stain was performed as described by W.H.O. (2008).

## Quantitative polymerase chain reaction (qPCR)

### qPCR on bacterial isolates derived from smears

The qPCR was performed on two different sample sets. First, cultured isolates from one of the two aliquots of the blood smears (506 isolates from 113 smears) were screened, targeting 5 genetic markers for *pagA* (BAPA), Ba-1, *lef*, GI4 and *capB* in a stepwise manner. All isolates were screened even if they were not phenotypically *B. anthracis.* The isolates were first screened with the SYBR Green assays using primers and targets in Table 1 as described by the manufacturer (CelGREEN, Celtic Molecular Diagnostics, Cape Town, South Africa). Isolates that were positive with SYBR Green (n=125) PCR assays on all the markers were then further confirmed using the TaqMan assay for targeting the Ba-1, *lef*, *capB, GI*4 (chromosomal) targets (Table 1) as described by Zincke et al. (2020) and the fluorescence resonance energy transfer (FRET) for BAPA (W.H.O., 2008). The inclusion of the chromosomal marker, Ba-1 and GI4 in the assay was based on the premise that it enhances the specificity of the assay, as detailed by (Ågren et al., 2013). The reaction mixtures for the SYBR Green assay, targeting the *pagA* (BAPA), Ba-1, *lef*, GI4, and *capB* primer sets, consisted of 0.5 µM of each primer. For Ba-1, *lef*, GI4, and *capB*, the mix included 1x SYBR Green (CelGREEN, Celtic Molecular Diagnostics, Cape Town, South Africa), while the BAPA assay utilized FastStart Essential Green Master (Roche, Basel, Switzerland). Each mixture also contained 5 µL of DNA, resulting in a total volume of 20 µL per reaction. Cycling conditions were: a pre-incubation at 95°C for 10 min (20°C/sec ramp), followed by 45 cycles of 95°C for 10 sec and 55°C for 20 sec (both at 20°C/sec ramp), then 72°C for 30 sec (20°C/sec ramp) with signal capture post-annealing. Denaturation involved an immediate 95°C step, cooling to 40°C for 30 sec (both at 20°C/sec ramp), then 80°C instantly with a 0.1°C/sec ramp for continuous signal reading. The process concluded with a cool-down to 40°C for 30 sec (20°C/sec ramp). The assay was performed using the QuantStudio 5 Real-time PCR system (Thermo Fisher Scientific, MA USA). Isolates that were positive for BAPA (n=14) were selected for further identification using gyrase B (*gyrB*) gene PCR as described by (Liu et al., 2021). We selected these isolates that were BAPA PCR positives as it has been hypothesised that closely related bacterial species might be responsible for the anti-PA serological reaction observed in anthrax nonendemic regions (Ochai et al., 2022). The cycle threshold (CT) cutoff was established at 35 for Ba-1 and GI4, as well as for *pagA* (BAPA), *lef*, and *capB* (Hassim, 2017b, Lekota et al., 2016, Zincke et al., 2020). For *B. anthracis*, we used *B. anthracis* Vollum strain as the positive control, and the 25 smear samples confirmed to be *B. anthracis* in the study of Hassim (2017a) were used as internal controls. We obtained a positive control (DNA from pure culture) for Bcbva from the Robert Kock Institute, Germany.

### qPCR from direct scrapings of blood smears

Secondly, the 1708 DNA samples obtained from blood smear scrapings were screened for the presence of the pXO1 plasmid, with qPCR assays targeting the *pagA* and *lef*, as well as pXO2 plasmid targeting *capB*. We also screened for the chromosomal markers Ba-1 of *B. anthracis* and GI4 region for Bcbva. To determine the presence of *pagA* (BAPA), qPCR was conducted using the FRET on the Light Cycler Nano (Roche, Basel, Switzerland). For the TaqMan assays, the reaction conditions were standardized to a 20-µL mixture containing 1 µL of the DNA template, 1x concentration of PrimeTime Gene Expression Master Mix (IDT, Coralville, IA, USA, Cat No. 1055772), along with primers and probes as listed in Table 3.

The thermal cycling conditions for TaqMan assays were set as follows: an initial denaturation at 95°C for 3 minutes, followed by 45 cycles of denaturation at 95°C for 20 sec and annealing/extension at 60°C for 30 sec. For *lef,* Ba-1, *capB* and *GI*4, the quantitative PCR TaqMan assay was performed using the QuantStudio 5 Real-time PCR system (Thermo Fisher Scientific, MA USA). Two duplex assays were created for the simultaneous detection of FAM– and VIC-labeled probes. The first duplex targeted Ba-1 and GI4 markers for species identification, while the second targeted *lef* and *capB* virulence markers from pXO1 and pXO2 plasmids, respectively. To prevent spectral overlap in the QuantStudio 5 instrument, color compensation was conducted with FAM and VIC probes, applying the results to duplex assay data. Tests included all 26 confirmed positive *B. anthracis* strains (∼1 ng DNA), and specificity checks involved DNA from Bcbva, and *B. cereus* ATCC 3999. The CT cutoff for positive samples was set at 35 for all the markers (Hassim, 2017b, Lekota et al., 2016).

### Conventional PCR

The 14 isolates from cultured blood smears that tested positive for pagA (BAPA) by the two qPCR methods described above were subjected to conventional PCR to allow for gel visualisation as described by Lekota et al. (2016). The reaction was set up with a total volume of 10 μL, containing 1x MyTaq PCR Master Mix (Bioline, MA, USA), 2 mM MgCl2, 0.2 µM of each *pagA* primer (Table 1), and 2 ng of the target DNA. The PCR amplification began with an initial denaturation step at 94°C for 5 min, followed by 35 cycles of denaturation at 94°C for 30 sec, annealing at 55°C for 30 sec. and extension at 72°C for 30 seconds, concluding with a final extension at 72°C for 5 min. The *B. anthracis* Vollum strain served as a positive control. *Bacillus cereus* ATCC3999 and distilled water served as negative controls, ensuring the assay’s specificity. Post-amplification, the PCR products were subjected to electrophoresis on a 3% ethidium bromide-stained agarose gel at 100 V for 90 min and visualized under UV light to confirm the presence of *Bacillus anthracis*-specific amplicons.

**Table 1:**
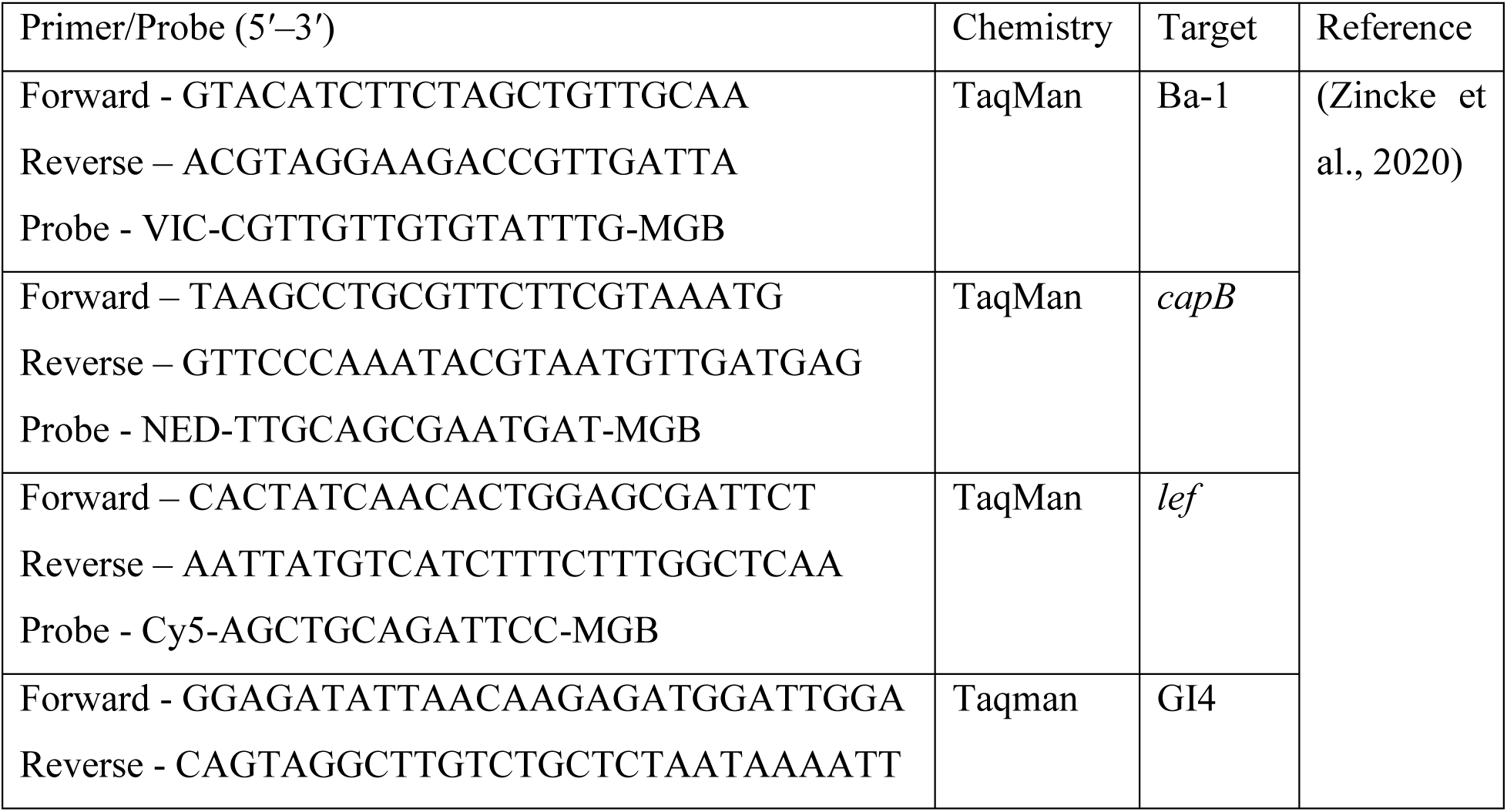
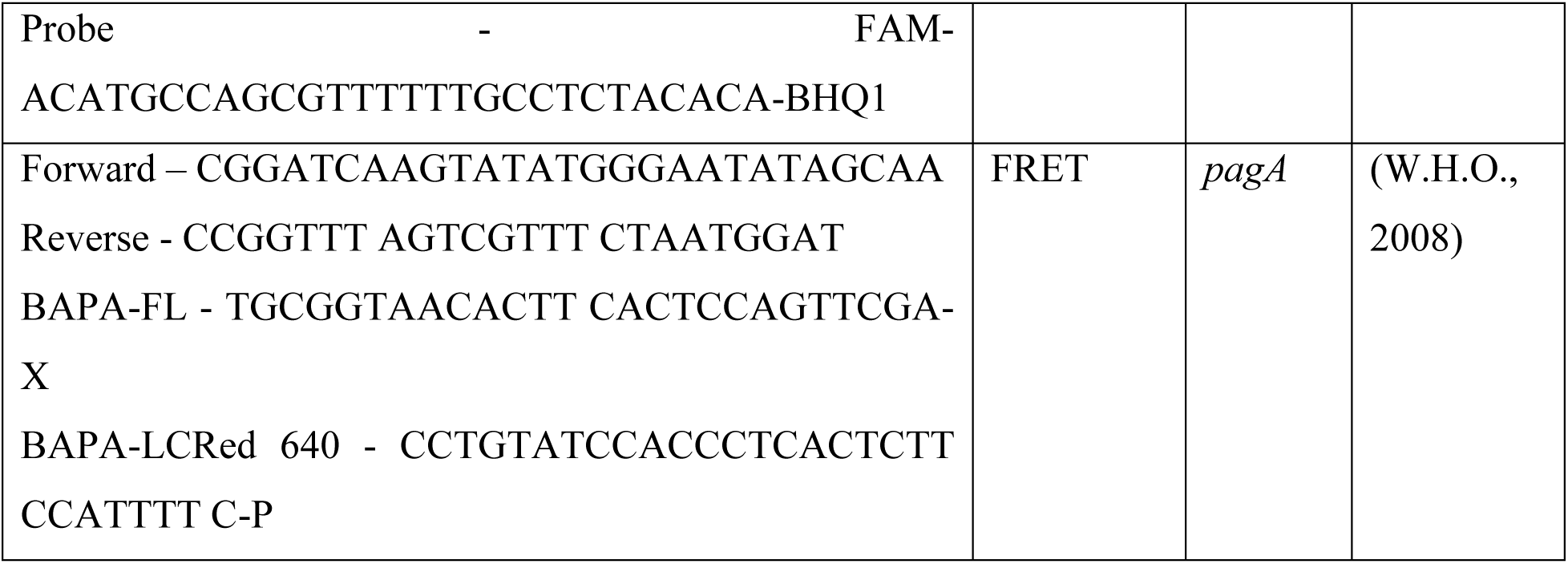
Primers, probes and gene targets for the detection of *Bacillus anthracis* from bacterial isolates cultured from blood smear samples from wildlife mortalities in Kruger National Park, South Africa by quantitative polymerase chain reaction (qPCR) assays. *Bacillus anthracis* protective antigen (BAPA), lethal factor (*lef*), chromosomal marker (Ba-1 and Genomic island 4: GI4) and the capsule region (*capB*) were used as molecular markers in this study. FRET stands for fluorescence resonance energy transfer.

### Molecular identification and phylogenetic analysis on bacterial isolates from smears

The 14 bacterial isolates from blood smear scrapings that tested positive for BAPA by the two qPCR approaches were subjected to additional molecular and phylogenetic analysisA. The gyrase B (*gyrB*) PCR product was sequenced for molecular taxonomic identification of the isolates. The PCR fragments of the *gyrB* gene of the selected isolates (n = 14), including 4 *B. anthracis* isolates based on microbiology (square-ended bacilli, colony morphology, penicillin and gamma phage sensitive), were sequenced at Inqaba Biotechnical Industries (Pty) Ltd., Pretoria, South Africa. A BLAST search query was performed to compare the *gyrB* nucleotide sequences from the *Bacillus* isolates with publicly available GenBank sequences in NCBI (http://www.ncbi.nlm.nih.gov; accessed on 08, March, 2023). Multiple sequence alignments of the mined *gyrB* reference sequences and *Bacillus* spp. strains sequenced in this study were performed using BioEdit 7 (Hall, 1999) and using the algorithm found in Clustal W MEGA11 as described by Tamura et al. (2021). With this alignment, we inferred the phylogenetic relationships of the *Bacillus* spp. isolates with respect to other related species and *B. anthracis.* The p-distance model was used to generate a neighbour-joining tree with 1000 bootstrapped replicates, using the MEGA 11.0 software (Tamura et al., 2021), and the phylogenetic tree was visualised using ITOL 5.0 (Letunic and Bork, 2021).

Positive PCR fragments targeting the *pagA* using the BAPA probe sequence (W.H.O., 2008) consisting of 240 bp were analyzed on the presumptive *Bacillus* spp. with sample numbers: AX2015 (1122; 1136; 1152; 1511 and 1277A) and AX2016 (1708NH and 1800) isolates and sequenced at Inqaba Biotechnical Industries (Pty) Ltd., Pretoria, South Africa. The BLASTn homology searches of the sequences were performed to assess homologous hits against the *pagA* region of the *B. anthracis* GenBank sequences available in NCBI (2023). Multiple sequence alignments of the *pagA* (BAPA) probe region were performed using BioEdit 7 (Hall, 1999). The isolates and/or PCR fragments that failed quality control (low-base calling during sequencing: the sequences where at least 90% of the nucleotides achieved a Phred score of less than 30) were excluded from this analysis.

## Data analysis

### Performance analysis of markers on bacterial isolates

All results for the qPCR of the isolates were presented as counts and percentages. To assess the performance of these molecular markers, we analysed 80 isolates that tested qPCR positive for individual markers or combinations of molecular markers using the probe-based approach. We used culture, microscopy, penicillin sensitivity, and gamma-phage sensitivity results as the gold standard (true positive/negative) for comparison with the assays (W.H.O., 2008). For the isolates that tested positive for any of the markers, we calculated the specificity, which detects true negative, and the sensitivity, which detects true positive (Griner et al., 1981). We also calculated the positive predictive value (PPV); probability that *B. anthracis* is present when the test is positive, the negative predictive value (NPV), probability that *B. anthracis* is absent when the test is negative, and the accuracy, which refers to the overall probability that a case is correctly classified (Griner et al., 1981). Results for specificity, sensitivity, and accuracy were presented in percentages and confidence intervals (CI) which are Clopper-Pearson CI (Clopper and Pearson, 1934) and the CI for the predictive values was calculated using the log method as described by Altman et al. (2000).

### Analysis of smears and direct qPCR of scrapings

The outcomes of the qPCR and microscopic examination of blood smears were represented as counts and percentages of positive samples. We evaluated the extent of agreement between the binary outcomes of the molecular tests and the results of the microscopic examination of the blood smears. This was done using a Cohen’s kappa (k) test (Cohen, 2003). For this analysis, kappa ≠ 0 implies that the extent of agreement between the two tests mentioned was significantly different from chance agreement. The measure of agreement was evaluated based on the criteria of Landis and Koch (1977), where <0 = poor; 0.01-0.20 = slight; 0.21-0.40 = fair; 0.41-0.60 = moderate; 0.61-0.80 =substantial; 0.81-1.00 = almost perfect.

Statistical analyses were conducted using R version 4.1.2 (R Core Team, 2021), and significance was evaluated with a threshold of alpha < 0.05.

## Results

### Isolation and identification of cultured samples

Out of the 1708 blood smear scrapings that were cultured, only 113 samples had bacterial growth from which a total of 506 pure colonies were isolated (some samples grew multiple different bacterial colonies). Only 4/506 colonies demonstrated morphological features that were consistent with those of *B. anthracis* (AX2015-1270, AX2015-1277A, AX2015-1152, and AX2015-1136). The colony morphology and structure of the four isolates on 5% SBA demonstrated non-heamolytic features, forming typical white-gray colonies with an oval, slightly granular appearance. The Gram-stained isolate smears from the 4/506 positive samples showed square-ended bacilli that are classical to *B. anthracis* (Figure 2). Upon examination of the polychrome methylene blue stained smears, the identified *B. anthracis* isolates appeared square-ended and encapsulated (with the exception of AX2015-1136, Figure 2G). The remaining 502 isolates from this study failed on all or some of the criteria (colony morphology, granularity or colour, hemolysis, capsule detection, penicillin and gammaphage sensitivity). The smear samples that failed to grow colonies included the known positive samples from which the 25 internal control samples were established, suggesting the *B. anthracis* endospores were no longer viable to establish culture from the smears.

**Figure 2:**
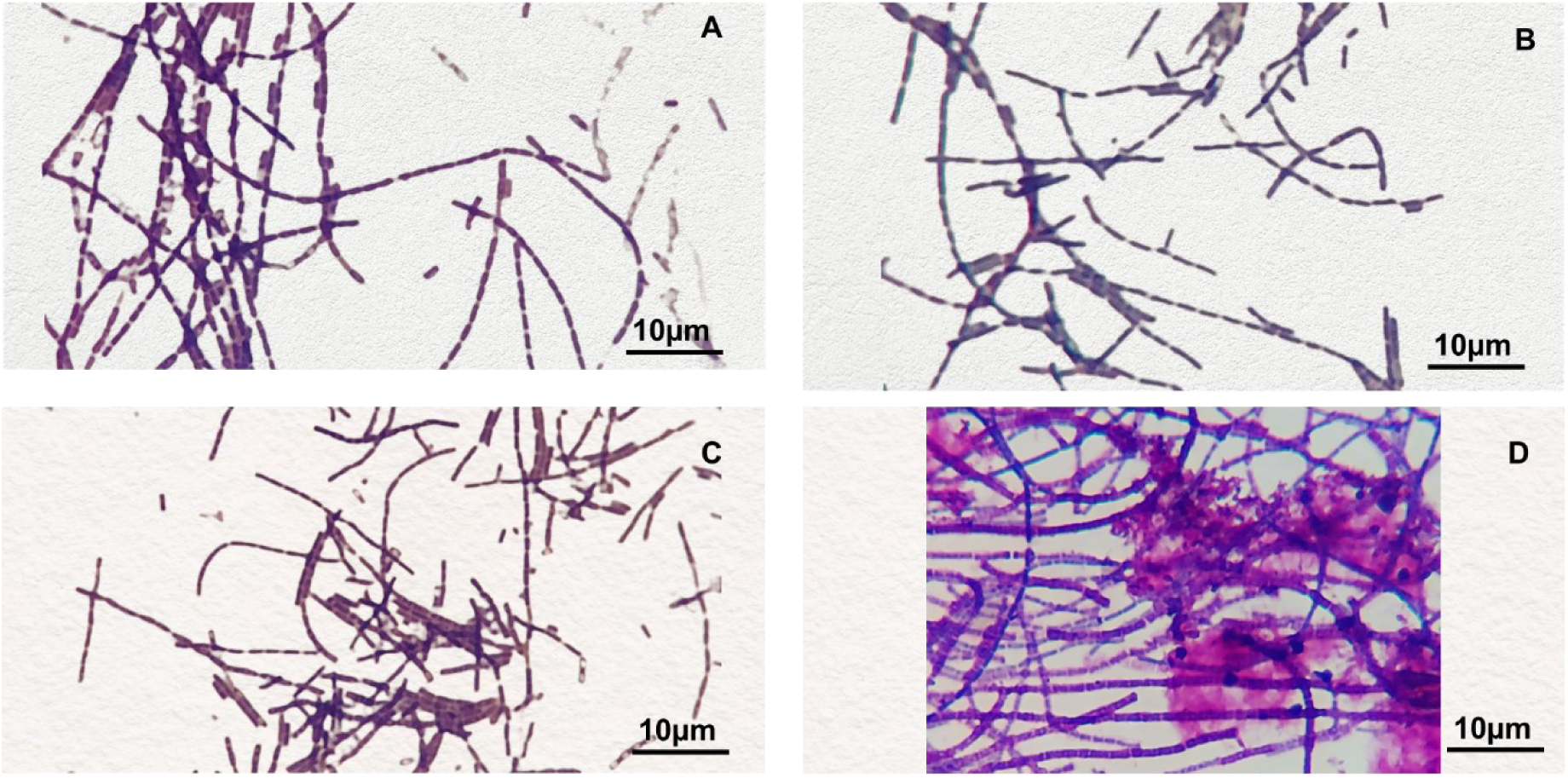
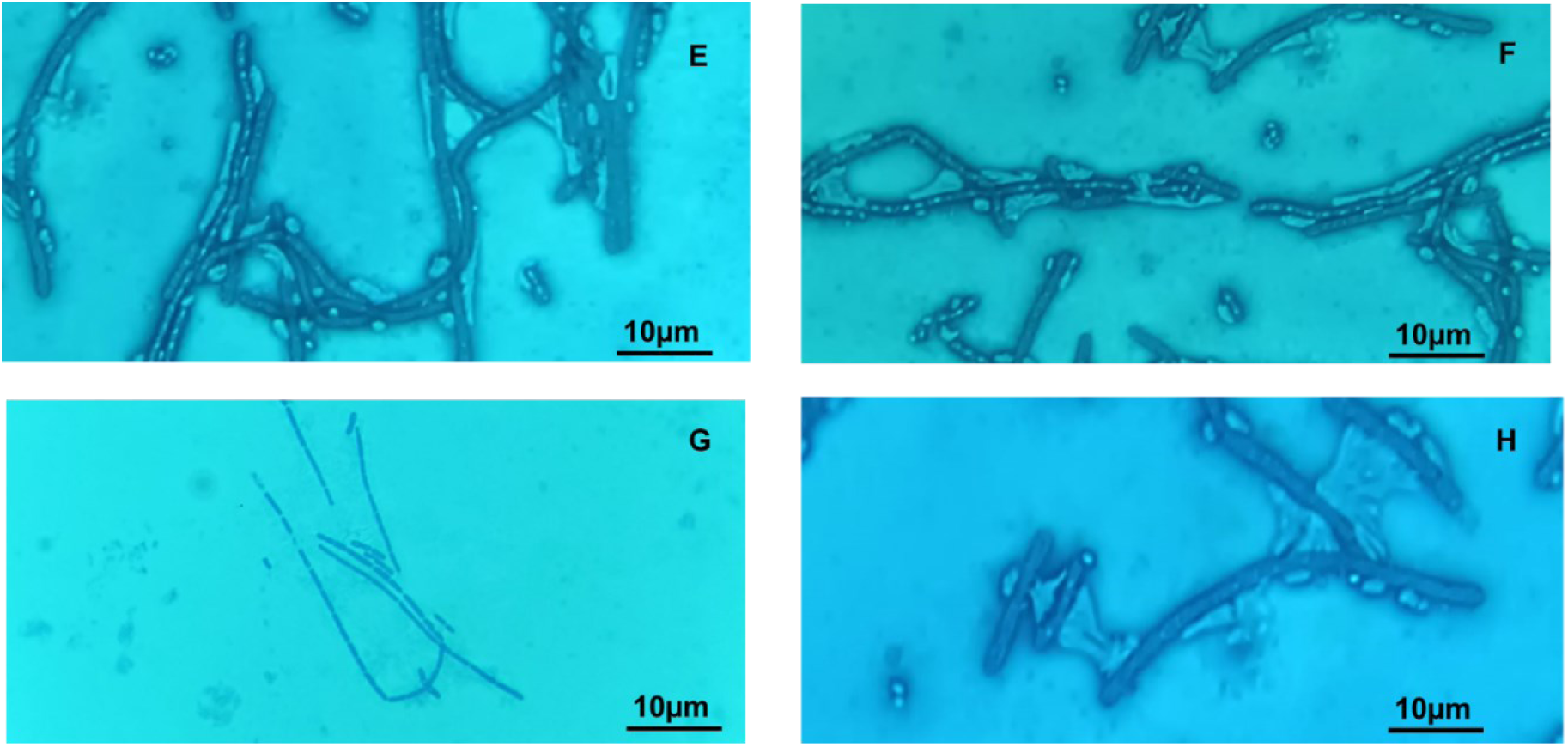
Microscopic examination of Gram and polychrome methylene blue stained cell cultures from bacterial isolates collected from blood smears of wildlife mortalities in Kruger National Park, South Africa that were identified as *Bacillus anthracis* through morphological methods. Images show square-ended bacilli: (A) Micrograph of isolate AX2015-1270, (B) Micrograph of AX2015-1152, (C) Micrograph of isolate AX2015-1136 and (D) Micrograph of isolate 1277A. Also, images show encapsulation of the bacilli: (E) Micrograph of isolate AX2015-1270, (F) Micrograph of AX2015-1152, (H) Micrograph of isolate AX2015-1277A and with the exception of (G) Micrograph of isolate AX205-1136 that demonstrated no capsule.

### Molecular analyses of bacterial isolates

Of the 506 bacterial isolates, only 125 (24.7%) tested positive for one or more of the molecular markers (Ba-1, *lef*, *pagA* (BAPA), *capB*) using SYBR Green (results not shown). Further confirmation of these 125 isolates using the probe-based approach showed that 80 isolates tested positive (Figure 3). The use of the “+” symbol in our context signifies the strategic combination of markers. The combination of BAPA + *lef* + Ba-1 successfully identified the four *B. anthracis* isolates that were confirmed through culture and microscopy. This combination yielded exclusively these four positives representing 5% (n=4) of the total.

**Figure 3:**
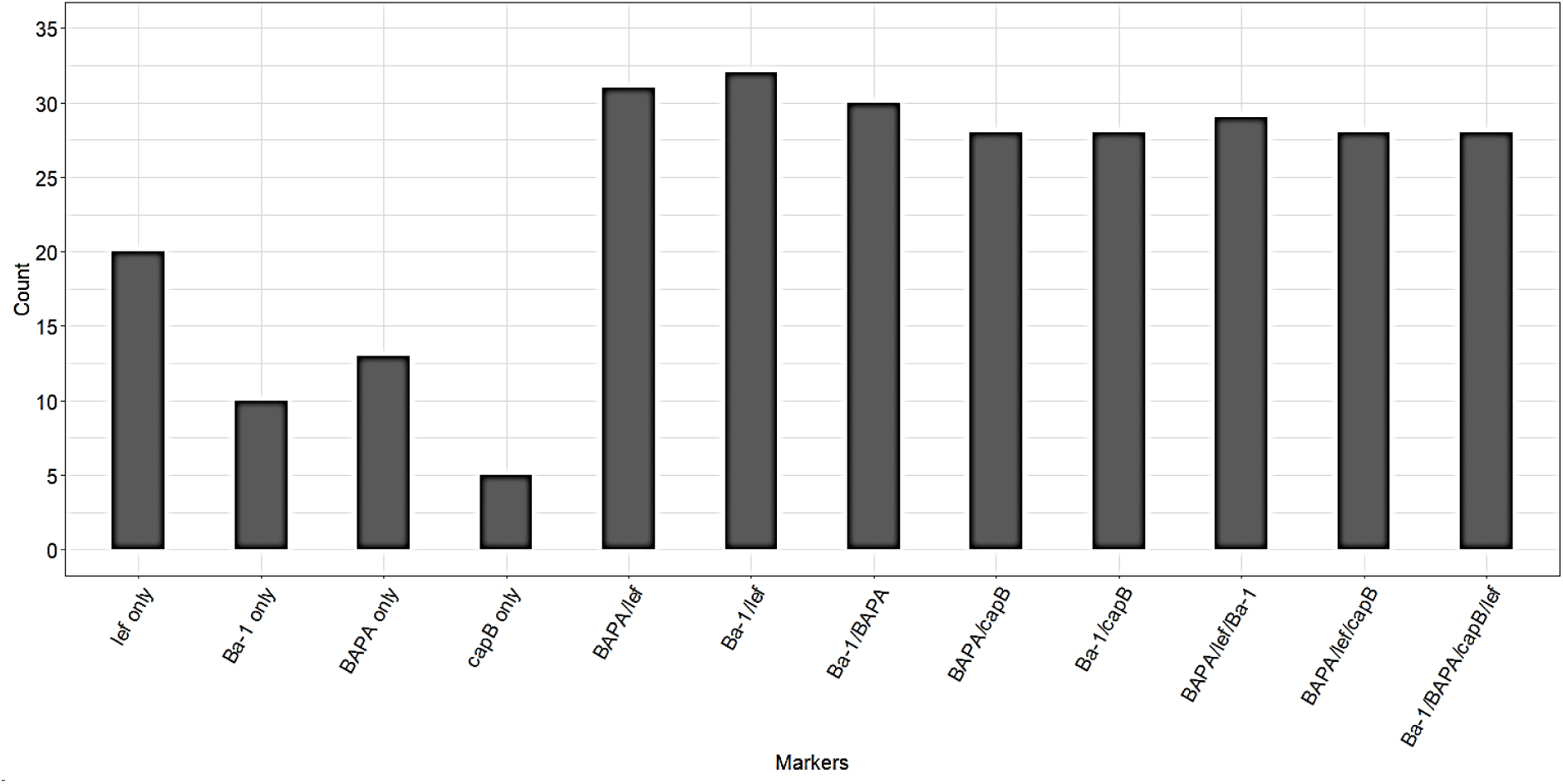
Bar plots showing counts of bacterial isolates grown from blood smears that tested positive for *Bacillus anthracis* based on different individual molecular markers or combinations of molecular markers. Bars show results of quantitative polymerase chain reaction (qPCR) of isolates positive using probe-based qPCR (total *n* = 80 positives). Samples were collected from blood smears of wildlife mortalities recorded from 2012-2020 in Kruger National Park, South Africa. *Bacillus anthracis* protective antigen (BAPA), lethal factor (*lef*), chromosomal marker (Ba-1) and the capsule region (*capB*) were used as molecular markers in this study. Results for GI4 were not included as all isolates including the internal controls tested negative. A total of 506 isolates were tested and only 80 isolates tested positive for individual markers or combinations of molecular markers using the probe-based approach.

### Microbiological screening of the 14 bacterial isolates that were confirmed to be positive on *pag*A (BAPA)

The most commonly used genetic marker for the diagnosis of *B. anthracis* is *pag*A (BAPA), and the use of this marker is recommended by the W.H.O. (2008); however, in our sample this marker was not specific to *B. anthracis*. Following a microbiologic screening of the 14 samples that tested positive for *pagA (*BAPA) by qPCR (Figure 4), 7/14 showed penicillin sensitivity, while only the samples that were identified as *B. anthracis* (i.e., by colony morphology, capsule staining, heamolysis and molecular markers) showed gamma phage sensitivity (Table 2). Only 2 of the 14 samples were haemolytic, and the *B. anthracis* strains were all non-hemaolytic (Table 2). The four *B. anthracis* strains were identified by qPCR markers Ba-1, *lef* and *capB* (although one tested negative for *capB*). Most marker combinations of chromosome and toxin genes as well as combinations of different toxin genes targets misclassified *B. anthracis.* A combination of microscopy and molecular techniques consisting of chromosome and toxin targets accurately detected *B. anthracis* while using capsule target underestimated *B. anthracis* due to the possible loss of the capsule encoding pXO2 (Figure 3).

**Figure 4:**
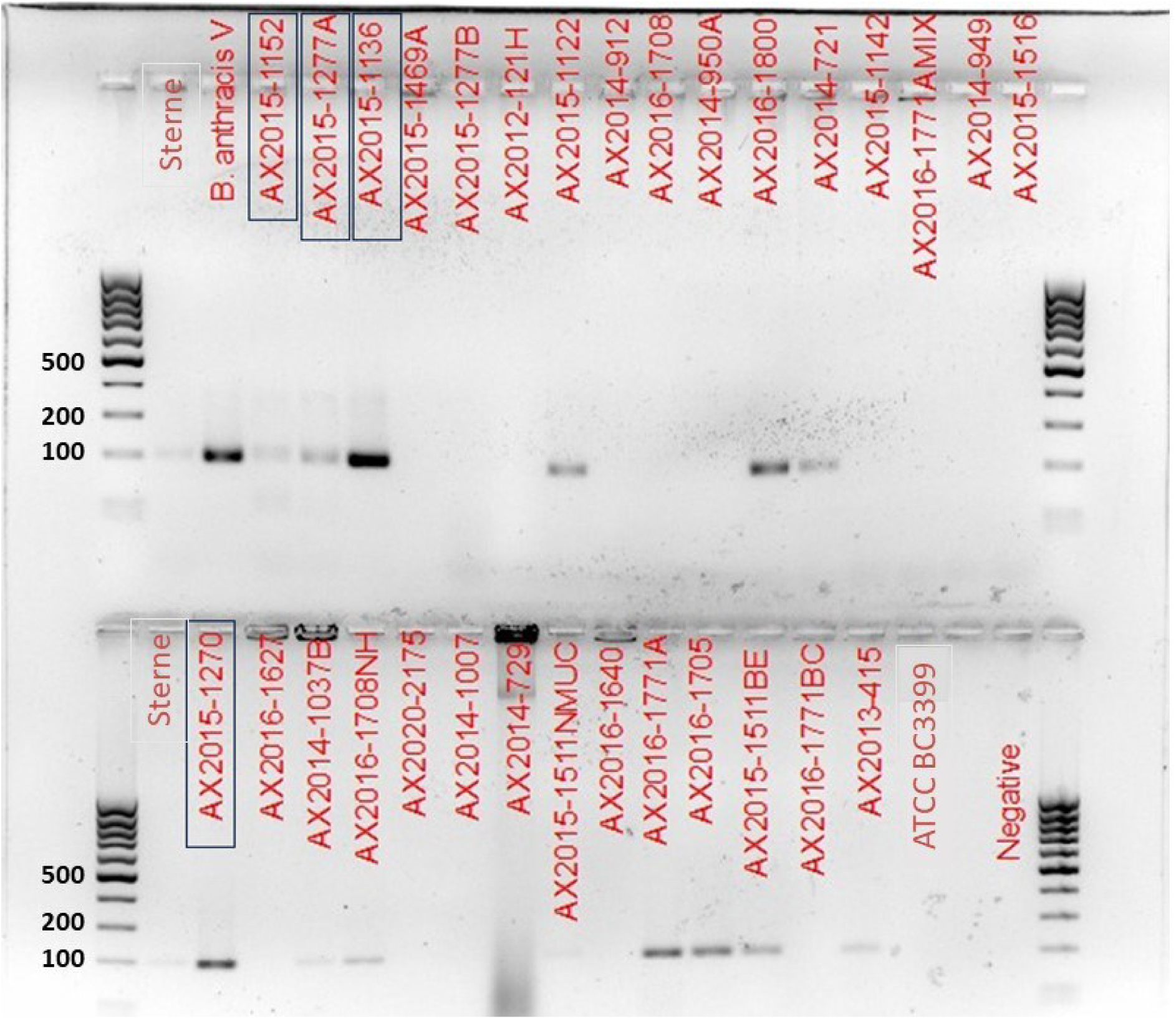
Agarose gel image of *Bacillus anthracis* protective antigen gene region, *pagA* (BAPA), for *Bacillus anthracis* and other bacterial species isolated from cultured blood smears obtained from wildlife mortalities in Kruger National Park, South Africa. The 100 bp (Thermo Scientific, MA, USA) ladder was used. The *B. anthracis* Sterne and Vollum (labelled as *B. anthracis* V) strains served as the positive controls. *Bacillus cereus* ATCC3999 and distilled water (labelled as Negative) were used as negative controls. Sample numbers highlighted with blue rectangles indicate *B. anthracis* confirmed samples. The assay was repeated three times.

**Table 2:** Results of penicillin and Gamma phage sensitivity, morphology and quantitative polymerase chain reaction (qPCR) for the Taqman probe-based chemistry using four different molecular markers (*Bacillus anthracis* protective antigen (BAPA), lethal factor (*lef*), and capsule (*capB*) genes and the chromosomal region (Ba-1)) for selected bacterial isolates collected from the scrapings of carcass blood smears from 2012-2015 from Kruger National Park, South Africa. These 14 isolates were those that tested positive for the *Bacillus anthracis* protective antigen BAPA marker. Genus identity was based on the *gyrase B* sequence data. The probe-based approach was only conducted on isolates that were positive with SYBR Green assay.

| Markers |  | Ba-1 |  | *lef* |  | BAPA |  | *capB* |  | Sensitivity and morphology |  |  |
| --- | --- | --- | --- | --- | --- | --- | --- | --- | --- | --- | --- | --- |
| Sample ID | Genus | SYBR<br>®<br>Green | Probe | SYBR<br>®<br>Green | Probe | SYBR<br>®<br>Green | Probe | SYBR<br>®<br>Green | Probe | Penicilli<br>n | Gammaph<br>age | Haemolys<br>is |
| AX2015-1122 | *Peribacillus* | + | - | - | + | + | + | - | - | + | - | - |
| AX2016-1800 | *Peribacillus* | + | + | - | + | + | + | - | - | + | - | - |
| AX2015-1277A* | *Bacillus* | + | + | + | + | + | + | + | + | + | + | - |
| AX2015-1136* | *Bacillus* | + | + | + | + | + | + | - | - | + | + | - |
| AX2015-1152* | *Bacillus* | + | + | + | + | + | + | + | + | + | + | - |
| AX2015-1270* | *Bacillus* | + | + | + | + | + | + | + | + | + | + | - |
| AX2016-1771A | *Bacillus* | - | - | + | + | + | + | - | - | - | - | + |
| AX2014-1037 | *Bacillus* | - | - | + | + | + | + | - | - | - | - | + |
| AX2016-1708NH1 | *Priestia* | + | + | - | - | + | + | - | - | + | - | - |
| AX2015-1511Nm | *Priestia* | + | + | - | + | + | + | - | - | - | - | - |
| AX2015-1511BE | *Priestia* | + | - | - | - | + | + | - | - | - | - | - |
| AX2016-1705 | *Priestia* | - | - | - | - | + | + | - | - | - | - | - |
| AX2013-415 | *Priestia* | + | - | + | - | + | + | - | - | - | - | - |
| AX2014-721 | *Priestia* | - | - | + | + | + | + | - | - | - | - | - |

Asterisk (\*) on sample IDs denotes *Bacillus anthracis*. The *Priestia* and *Peribacillus* species were previously identified as *Bacillus*: however, Bergey's manual (Bergey, 1923) for the nomenclature of bacteria still refers to these as *Bacillus*.

### Performance of the pXO1, pXO2 gene and chromosomal markers

The different molecular markers alone and in combination demonstrated varying specificity, sensitivity, PPV, NPV, and accuracy (Table 3). The 80 isolates identified as positive for BAPA by the qPCR/probe approach in this study include the 4 *B. anthracis* isolates identified from the smears. The *lef* marker demonstrated the lowest specificity and accuracy (51.2% and 72.5%, respectively; Table 3). Specificity and accuracy for Ba-1, BAPA, and *capB*, for qPCR were all above 60.0%, with Ba-1 having the lowest and capB having the highest specificity and accuracy (Table 3). The combination of markers increased the specificity and accuracy of these markers. Combinations of Ba-1+lef, BAPA+*lef,* and Ba-1+BAPA showed specificities and accuracies of over 95% (Table 3). The specificity and accuracy were 100% and 98.8%, respectively, for all combinations of Ba-1+*capB*, BAPA+*capB*, Ba-1+BAPA+*capB*+*lef*, and BAPA+*lef*+*capB*, however, with a sensitivity of 96.55 % (Table 3). The combination of BAPA+*lef*+Ba-1 showed a specificity, sensitivity, and accuracy of 100% which is the overall probability that a case is correctly classified.

**Table 3:** Performance of quantitative polymerase chain reaction (qPCR) probe-based diagnostic assays for detecting *Bacillus anthracis,* using individual markers as well as combinations of markers as assessed by their specificity, sensitivity, positive predictive value, negative predictive value and accuracy. All results are shown in percentages with confidence intervals (CI; 95%) in parentheses. The gold standard assessment of a true positive used in this analysis was culture identification, microscopy, and penicillin and Gamma phage sensitivity. Samples used here (n=80) include *Bacillus anthracis* (Ba) and other bacterial species isolated from cultured blood smears obtained from wildlife mortalities in Kruger National Park, South Africa. *Bacillus anthracis* protective antigen (BAPA), lethal factor (*lef*), chromosomal marker (Ba-1) and the capsule region (*capB*) were used as molecular markers in this study.

| Markers | Specificity (CI) % | Sensitivity (CI) % | Positive predictive value(CI) % | Negative predictive value (CI) % | Accuracy (CI) % |
| --- | --- | --- | --- | --- | --- |
| Ba-1 only | 75.61(59.70-87.64) | 72.50 (56.11-85.40) | 74.36 (62.46-83.49) | 73.81 (62.33-82.76) | 74.07 (63.14-83.18) |
| *lef* only | 51.22(35.13-67.12) | 72.50 (56.11-85.40) | 59.18(50.75-67.11) | 65.62 (51.54-77.41) | 75.00 (64.06-84.01) |
| BAPA only | 67.50 (50.87-81.43) | 72.50 (56.11-85.40) | 69.05 (57.85-78.38) | 71.05 (58.68-80.93) | 70.00 (58.72-79.74) |
| *capB* only | 90.00 (78.19-96.67) | 96.67 (82.78-99.92) | 85.29 (71.58-93.03) | 97.83 (86.73-99.68) | 92.50 (84.39-97.20) |
| Ba-1 + *lef* | 94.12 (83.76-98.77) | 100.00 (88.06-100.00) | 90.62 (76.33-96.66) | 100.00 (92.60-100.00) | 96.25 (89.43-99.22) |
| BAPA + *lef* | 96.08 (86.54-99.52) | 100.00 (88.06-100.00) | 93.55 (78.85-98.26) | 100.00 (92.75-100.00) | 97.50 (91.26-99.70) |
| Ba-1 + *capB* | 100 (93.02-100) | 96.55 (82.24-99.91) | 100.00 (87.66-100.00) | 98.08 (88.14-99.72) | 98.75 (93.23-99.97) |
| BAPA + *capB* | 100 (93.02-100) | 96.55 (82.24-99.91) | 100.00 (87.66-100.00) | 98.08 (88.14-99.72) | 98.75 (93.23-99.97) |
| Ba-1 + BAPA + *capB* + *lef* | 100 (93.02-100) | 96.55 (82.24-99.91) | 100.00 (87.66-100.00) | 98.08 (88.14-99.72) | 98.75 (93.23-99.97) |
| BAPA + *lef* + *capB* | 100 (93.02-100) | 96.55 (82.24-99.91) | 100.00 (87.66-100.00) | 98.08 (88.14-99.72) | 98.75 (93.23-99.97) |
| BAPA + *lef* + Ba-1 | 100.00 (93.02-100.00) | 100.00 (88.06-100.00) | 100.00 (88.06-100.00) | 100.00 (93.02-100.00) | 100.00 (95.49-100.00) |
| Ba-1 + BAPA | 98.04 (89.55-99.95) | 100.00 (88.06-100.00) | 96.67 (80.64-99.51) | 100.00 (92.89-100.00) | 98.75 (93.23-99.97) |

### *Bacillus* spp. differentiation using *gyrB*

The BLASTn identification of the *gyrB* gene from the 14 selected bacterial isolates (i.e., those positive for BAPA) and subsequent phylogenetic analyses identified three genetic clusters, *B. cereus sensu lato* (comprising of *B. cereus* and *B. anthracis* found in this study), *Peribacillus* spp. and *Priestia* spp. (Figure 5). The latter two clusters were previously part of *Bacillus* and are recently proposed new genera (Gupta et al., 2020) but are still documented as *Bacillus* spp. according to the Bergey’s’s manual (Bergey, 1923). The AX2015 strains (1152, 1277A, 1270 and 1136) grouped in the *B. cereus sensu lato* cluster with reference isolates *B. anthracis* (FDAARGOS 695 and Kanchipuram) as the closest related strains. The isolated AX2016-1771A strain clustered with *B. anthracis*, and also within a cluster including atypical *B. cereus*, although it had phenotypic characteristics with *B. cereus* as it was classified as haemolytic. AX2014-1037B; AX2015-1122 and AX2016-1800 grouped in the *Peribacillus* cluster (Figure 5). AX2015-1511BE grouped with *Priestia megaterium* reference strains, and AX2016-1708NH1 grouped closely with the *Priestia aryabhattai* reference strains (Figure 5). The following isolates AX2013-415, AX2014-721, AX2015-1511 Nm and AX2016-1705 were excluded from the tree as they failed to pass the quality control.

**Figure 5:**
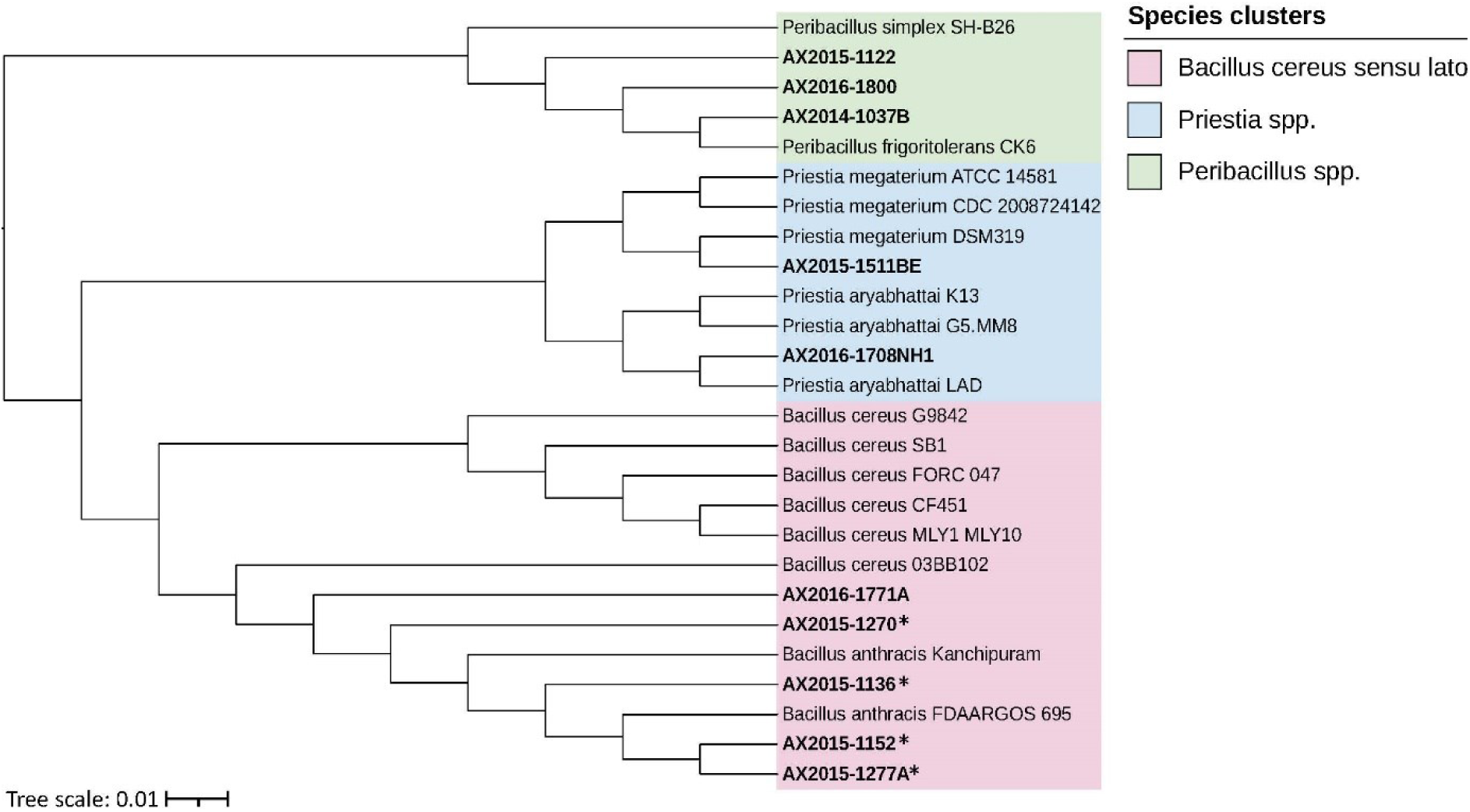
Phylogenetic tree of bacterial isolates from Kruger National Park, South Africa based on the *gyrase B* gene constructed using the neighbour-joining method and p-distance model among three closely related genera (all formerly *Bacillus* spp.). Isolates starting with AX are from this study and were matched against the closest National Center for Biotechnology Information (NCBI) reference isolates (through BLASTn searches) of *Bacillus cereus sensu lato*, *Priestia* spp., and *Peribacillus* spp. The scale bar represents 0.010 substitutions per nucleotide position. All positions containing gaps and missing data were eliminated from the dataset (complete deletion option). Isolates classified/identified as *B. anthracis* from this study following confirmation with microscopy, culture, molecular diagnosis, and penicillin and gammaphage sensitivity are denoted with an asterisk (*).

### The *pagA (*BAPA) sequence alignment of the selected isolates

The 240 bp *pag*A (BAPA) region of AX2014-721, AX2015 (1122; 1136; 1152;1277A) and AX2016 (1771A; 1705; 1800; 1708NH1) were aligned against the NCBI reference strain of *B. anthracis* DFRL BHE-12 *pagA* gene region and the BAPA probes (See Figure 6 with probe sequence). The results showed no difference in comparison to the reference *B. anthracis* strains and showed the *pagA* region of the probe was completely conserved across the isolates (Figure 6).

**Figure 6:**
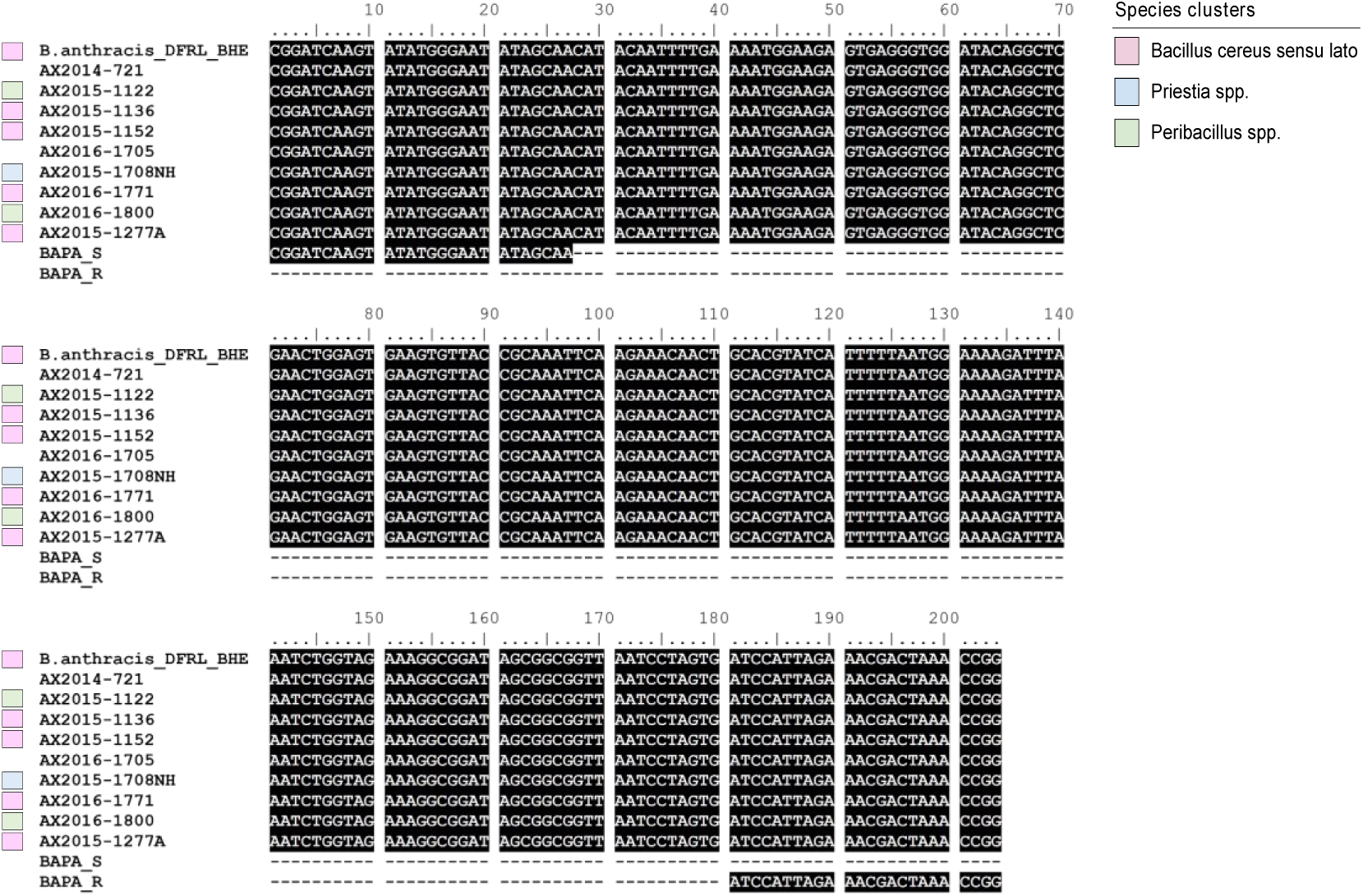
Multiple nucleotide sequence alignment of the *Bacillus anthracis* protective antigen (BAPA) probe region (targeting the *pagA* gene of *B. anthracis*) of isolates obtained from this study (starting with AX) versus the NCBI reference strain *B. anthracis* DFRL BHE-12. BAPA_S (Forward) and BAPA_R (Reverse) represent the BAPA probe sequences. Coloured blocks represent the related species clusters in which the isolates grouped as shown in Figure 4. The nucleotide sequences were aligned using MAFFT V7.0 (Katoh and Standley, 2013). Two samples (AX2014-721 and AX 2016-1705) are missing colour blocks as their species’ clusters were not assessed in the previous analysis (Figure 5).

### Probe-based qPCR and microscopy results of scraped blood smears

The DNA directly extracted from blood smears (n = 1708 samples) revealed 890 samples that tested positive for at least one of the molecular markers evaluated (Table S1). However, of these 890 samples, 165 (18.5%) tested positive for *lef* only and 112 (12.6%) for *pag*A (BAPA) only, and not for any other marker. Details of other markers and combinations of markers are found in Table S2. A combination of Ba-1 + BAPA + *cap*B + *lef* yielded a total of 393 positive samples (44.2% of the 890 samples; Table S1).

Microscopic evaluation of the 1708 blood smears detected 24.9% (425) of positive samples based on presence of bamboo-shaped, square-ended bacilli. However, when combining molecular and microscopy results, BAPA + *lef* + microscopy yielded 395 positives (23.1% of 1708 samples), Ba-1 + BAPA + microscopy had 398 positives (23.3% of 1708 samples), BAPA + *cap*B + microscopy had 400 positives (44.9% of 890 samples), *lef* + *capB* + microscopy had 401 positives (23.4% of 1708 samples), Ba-1 + *cap*B + microscopy had 397 positives (23.2% of 1708 samples), Ba-1 + BAPA + *lef* + microscopy had 393 positives (23.00% of 1708 samples), while Ba-1 + BAPA *+ lef* + *capB* + microscopy yielded 391 positives (22.9% of 1708 samples). All combinations included the 25 internal controls and the four *B. anthracis* isolates from this study except for AX2015-1136 which was missing in the combinations including *capB*.

There was a significant and moderate agreement between the binary outcomes of the molecular tests (combining Ba-1 + BAPA + *lef + capB*) and the results of the microscopic examination of the blood smears (kappa = 0.73, 95% CI: 0.67-0.78, *p*<0.0001). All samples were negative for the genomic island GI4 of Bcbva.

## Discussion

In this study, we investigated if bacteria closely related to *B. anthracis* can complicate anthrax surveillance and diagnostics based on molecular markers from wildlife mortalities in Kruger National Park (KNP), South Africa. Based on variation in the *gyrB* gene, we showed that blood smears can contain bacteria from *Priestia* spp., *Peribacillus* spp., (both former *Bacillus* spp.) and *B. cereus sensu stricto*, that cross-react with common molecular markers used for *B. anthracis*. Specifically, they cross-react with *pagA* (BAPA) or *lef* that have been used as the antigen target in anthrax serology (Brézillon et al., 2015, Marston et al., 2016). These closely related bacteria could be pathogenic, commensal, or environmental contaminants. Therefore, it is important to consider the potential that molecular markers to detect pathogenic organisms may risk cross-reacting with closely related organisms that co-occur with the pathogenic organism, and evaluate the use of a combination of different markers as done in this study. Lastly, we found good agreement between anthrax diagnoses based on microscopy of blood smears and molecular techniques using a series of markers. This indicates that such methods can accurately identify anthrax in diagnostic samples, which might render the conventional gold standard culture confirmation unnecessary. This shift could reduce the biosafety and biosecurity risks associated with the traditional approach, which are greater than those posed by these proposed techniques.

The identification of species closely related to *B. anthracis* on diagnostic blood smears can complicate anthrax diagnosis as these species may share similar genetic markers with *B. anthracis*, leading to false positive results from molecular diagnostics. In addition, other genera, such as *Peribacillus* and *Priestia* (Loong et al., 2017, Lekota et al., 2018), can also complicate anthrax diagnosis. These species may not share as many genetic markers with *B. anthracis* as the *B. cereus* group, but still have some similarities that could lead to false positive results as seen in this study when performing qPCR diagnostics using only *lef*, Ba-1, or *pagA* (BAPA) markers (Figure 6). For instance, *Peribacillus* and *Priestia* genera have been reported to have similar 16S rRNA gene sequences and protein profiles as *Bacillus* (Bhattacharjee et al., 2023), which can lead to misidentification. This suggests the presence of other bacterial species that share similar *pagA* to that of *B. anthracis* with significant implications for *pagA*-based ELISA. The results of our study show that other closely related organisms can react to *pagA* and produce false positive results as hypothesised in a serological study conducted in KNP (Ochai et al., 2022). It is therefore necessary to consider using other genetic markers or a combination of markers to confirm the presence of *B. anthracis*.

Specifically, Lekota et al. (2018) demonstrated that the capsular region or polyglutamate regions (*capABC*) are the ones that complicate anthrax diagnosis. Lekota et al. (2018) reported that *capC* is not specific to *B. anthracis*. Thus, combining capsule markers such as the *capB* in this study with other markers increased the specificity. However, in this study, *capB* had a lower sensitivity (96.67%) that should be interpreted with caution owing to the small number of samples that were confirmed as *B. anthracis*. Because the virulence factors of *B. anthracis* occur in closely related *Bacillus* species (Baldwin, 2020), the combination of chromosome, toxin and capsule genes may yield the best diagnostic result as seen in this study. The BAPA + *lef* + Ba-1 combination showed 100% specificity, sensitivity, and accuracy.

Archival smears are a useful resource for retrospective studies and retrieval of environmentally persistent pathogens like *B. anthracis* (Vince et al., 1998). We were only able to culture *B. anthracis*, as defined by microscopy, culture, molecular diagnosis, and sensitivity to penicillin and gammaphage, from four samples collected during the 2015 outbreak from impala (*Aepyceros melampus*). These samples did not include any of the 25 *B. anthracis* internal controls collected in 2012-2013. This indicates that the endospores were not viable after 10 years from these 25 smears, which were known to be *B. anthracis* cases from previous work. This agrees with the findings of Hassim (2017b) who reported that the longer a smear is stored, the harder it is to recover *B. anthracis,* and this may also affect the quality of the DNA extracted from such samples. The capsule found on the pXO2 plasmid was potentially missing for 1 of the 4 *B. anthracis* isolates obtained from the smears. This has previously been reported to occur in the long term storage of isolates (Marston et al., 2005). The mechanism of how the plasmids are lost is still not properly understood but it is hypothesised to be due to damage to the DNA or following nutrient deficiency over time (Marston et al., 2005). This suggests the possibility that archival smears might benefit from storage in climate-controlled conditions to prolong their shelf life. Additionally, although *B. anthracis* can survive for extended periods, it doesn’t guarantee it always will, indicating that storage conditions warrant further evaluation.

The chromosomal marker Ba-1 has been reported to be very specific to *B. anthracis* (Zincke et al. (2020). However, in this study, of the isolates that tested positive for Ba-1, only 4/42 were confirmed to be *B. anthracis* based on morphological, microscopic and sensitivity tests (gamma-phage and penicillin). The difference between our study and Zincke et al. (2020) might be due to degradation of the samples used in this study being archived over time. It may also be due to the different samples pools, where Zincke et al. (2020) evaluated the Ba-1 marker using samples of Bcbva, *B. cereus*, and *B. thuringiensis,* whereas the majority of the bacteria isolated in this study were *Priestia* spp, and *Peribacillus* spp. It is known that *Priestia* spp. and *Peribacillus* spp. are quite ubiquitous as they can be found in soil, faeces and the plant rhizosphere (Gupta et al., 2020), complicating anthrax diagnosis. All the samples from this study were negative for GI4, and there have been no reports of Bcbva outside of west Africa; thus, Bcbva may not be present in KNP, South Africa, and the GI4 may not be a viable marker for pathogenic strains in southern Africa. However, it is still important to consider GI4 screening for Bcbva. The ecological range of Bcbva is not fully understood, especially its presence in transitional areas between humid forests and dry savannas, which are typical habitats for *B. anthracis*. It is therefore important to investigate other non-traditional regions, using new diagnostic tools like Bcbva-specific proteins for detection. Understanding Bcbva’s distribution is crucial for assessing risks in these regions and guiding future surveillance and research efforts. This suggests that geographic region-specific diagnostics could be developed in the identification of anthrax-like cases, if these rare cases do exist.

From this study, accurate detection of *B. anthracis* in a sample may be improved with a stepwise approach based on multiple genetic markers, especially in situations where culture is not possible. Ba-1 has been used by Blackburn et al. (2014) and Zincke et al. (2020) in the diagnosis of *B. anthracis*. However, both studies performed additional MLVA-based or WGS based efforts to confirm species or further characterize them which eliminated the possibility of an overestimation. In our study we demonstrated that using only the Ba-1 or *lef* marker produced non-specific diagnostic results, with specificities estimated as 92.1% and 79.0% for the probe-based assay, respectively. However, testing for the presence of both markers decreased the number of false positives, with a specificity 96.1% (Table 3). More precise results were obtained while testing for the presence of BAPA *+ capB,* Ba*-*1 *+ capB*, BAPA *+ lef + capB*, and Ba-1 *+* BAPA *+ lef + capB*, with all combinations having a specificity of 100%, sensitivity of 75.0%, and accuracy of 98.8%. However, the problem with these combinations is that there is a high chance of misdiagnosing *B. anthracis* isolates that are missing the capsule, as was the case of AX2015-1136. The only combination that demonstrated a specificity, sensitivity and accuracy of 100% was BAPA *+ lef +* Ba-1 (Table 3). Therefore, the combination of BAPA *+ lef +* Ba-1 could be a useful molecular strategy to diagnose *B. anthracis* in the absence of other methods like culture and microscopy. However, the addition of *capB* is always important in identifying capsule producing *B. anthracis*. It is however important that a negative *capB* result might stem from factors such as insufficient amplification prior to the cycle threshold or DNA degradation. Using MLVA and genotyping, pXO2 samples are more easily identified (Mullins et al., 2013, Easterday et al., 2005), where these samples are integrated into phylogenetic trees alongside strains verified via qPCR, regardless of the absence of *capB*. Although such instances in the literature are not widespread and warrant further examination.

Of the 890 samples screened directly from the blood smear that tested positive based on at least one probe-based qPCR, 15.2% tested positive for only *lef* and negative for other markers and microscopy. The absence of other genetic markers and the negative result on microscopic examination suggests that *lef* is also non-specific to *B. anthracis,* present in other species. In this study, *lef* appears less specific to *B. anthracis* as compared to BAPA or other gene markers used in the diagnosis of anthrax. This report is consistent with the work of Zincke et al. (2020) who demonstrated that *lef* could also amplify *B. thuringiensis* serovar Kurstaki HD1 and *B. cereus* G9241 that also possesses a pXO1-like plasmid that carries the genes for anthrax toxin subunits. The presence of *lef* in non-*B. anthracis* pathogenic *B. cereus* has also been reported in humans (Hoffmaster et al., 2004); incorporating other markers or techniques when testing for the presence of *B. anthracis* in a sample could improve diagnosis. The combination of markers from the two plasmids or from the plasmids and the chromosome greatly improved the diagnosis of anthrax with fewer false positive results. The addition of microscopy to the various combinations increased the accuracy of the results significantly as there was very little variation on the number of positive samples when comparing all the combinations with microscopy. Also, the combination of microscopy and the genetic markers can accurately diagnose *B. anthracis* hence reducing the need for culture and proliferation. We also found that *lef* was less specific on the isolates, as compared to BAPA and *capB*. Penicillin sensitivity was also observed in two of the non-*anthracis* isolates; however, sensitivity to gamma phage was only seen in the *B. anthracis* isolates. The combination of Ba-1, BAPA, *capB* and *lef* was found only in the *B. anthracis* isolates with the exception of AX2015-1136, which was missing the capsule. Furthermore, the sample should test positive for BAPA+*lef*+Ba-1 or any of the combinations containing *cap*B (such as BAPA+*cap*B, Ba-1+*capB*, BAPA+*lef+cap*B, Ba-1+BAPA+*cap*B+*lef)*.

The significant agreement between the microscopic and molecular diagnosis of anthrax in this study demonstrates the usefulness of the microscopic technique in field and onsite diagnosis of the pathogen. The combination of microscopy with qPCR, using DNA extracted from blood smear scrapings, marks an advancement in diagnosing *B. anthracis*. This approach could significantly lessen the dependency on conventional culture-based diagnostic methods. Furthermore, it becomes especially critical in the context of increasing bioterrorism threats (Binkley et al., 2002), offering a rapid, specific, and safer alternative for identifying *B. anthracis*. Microscopy allows for the initial, rapid identification of the characteristic *Bacillus* rods, which may be indicative of *B. anthracis*, particularly when observed in the context of a clinical presentation consistent with anthrax. However, microscopy alone cannot provide a definitive diagnosis due to the presence of morphologically similar *Bacillus* species. This is where qPCR becomes invaluable; by targeting specific DNA sequences which together can be unique to *B. anthracis*, qPCR offers a highly sensitive and specific identification method. When applied to DNA extracted directly from blood smear scrapings, these two techniques can rapidly confirm the presence of *B. anthracis*, circumventing the time-consuming and potentially hazardous process of culture (Binkley et al., 2002). This dual approach not only enhances diagnostic speed and safety but also supports early intervention and effective public health responses to anthrax outbreaks (Sabra et al., 2023).

Microscopy has been a traditional and valuable tool in the diagnosis of anthrax. In this study, 24.9% (425) of the blood smears which included the 25 internal controls were positive for anthrax under the microscope, while an average of 23.1% were positive for combinations of markers and microscopy. This shows that molecular tests should not entirely replace microscopy as a diagnostic technique for anthrax. A study comparing the performance of PCR and microscopy in diagnosing cutaneous anthrax found that PCR had a higher sensitivity (100.0%) compared to microscopy (60.0%) (Berg et al., 2006). However, the authors noted that microscopy can still provide useful information on the clinical presentation and progression of the disease as a diagnostic tool. Similar studies have reported on the usefulness of microscopy as viable and important in resource-poor or field conditions using different criteria (Aminu et al., 2020). The importance of combining different methods can not be over emphasised especially with recent reports of Bcbva possessing several characteristics of *B. anthracis* (Klee et al., 2006, Antonation et al., 2016). For example, organisms are non-hemolytic and both form rods in chains that can be difficult to differentiate as reported by Antonation et al. (2016). With ongoing advancements in next-generation sequencing and decreasing costs, leveraging computational methods with robust bioinformatics can significantly contribute to achieving a robust diagnostic of anthrax and successful differentiation from Bcbva and other anthrax-like pathogens. The findings of this study offer substantial implications for public health and One Health initiatives (Bhattacharya et al., 2021). These insights contribute to advancing public health and One Health gains by providing more accurate, efficient, and accessible diagnostic approaches for anthrax detection, ultimately aiding in the prevention and control of the disease in livestock, wildlife and human populations.

### Limitations of the Study

1. Despite identifying *B. anthracis* in smears from 2012-2015 outbreaks, culture success from these archived slides was limited to the 2015 outbreak, possibly due to challenges in the viability of *B. anthracis* endospores in blood smears over time.
2. The storage of the blood smears over time could have had an impact on the quality of the smears and could introduce possible bacterial contamination.
3. The determination of sensitivity, specificity, and accuracy were based on only 4 positive samples plus 25 internal controls. Therefore, assessing the performance of the assays on a larger number of samples that includes more culture/gold standard-confirmed positive cases would be beneficial.

## Conclusion

Results of this study demonstrate that diagnostic markers and techniques that are specific to *B. anthracis* could reduce the complications in detection that are currently experienced, especially with an increase in the exploration of the potential sharing of genetic material amongst the *B. cereus sensu lato* members. Microscopy remains a very valuable tool in confirming the presence of *B. anthracis* in the field and in resource-limited settings, as well as a confirmatory tool. Accurate diagnosis with microscopy and combination of markers can reduce or eliminate the need for culture and bacterial proliferation. The presence of non-*B. anthracis* organisms harbouring similar genes may complicate anthrax diagnosis in the field. Lastly, the study identifies that the combination of BAPA+*lef*+Ba-1 yields the most specific, sensitive, and accurate results. Additionally, employing combinations with BAPA+*lef*+*capB*, with microscopic analysis, can enhance diagnostic confirmation and reduce false positives and reduce the need for culture, as revealed in this research.

## Suggestions for Future Research

Future studies to develop gene markers that better differentiate *B. anthracis* from its close neighbors in southern Africa may benefit the diagnosis of anthrax in this region. Whole genome analysis looking at the presence of these genes in non-*anthracis* species could inform selection of the best regions to target for primer and probe development. Studies evaluating bacterial isolates from different geographical regions could be informative to test for the specificity of developed markers for anthrax diagnosis and reduce or eliminate cross reactivity. Lastly, exploring the diversity of non-pathogenic microbes related to pathogenic ones can provide more information about the potential for the emergence of novel pathogenic microbes.

## Data Availability Statement

At the time of publication, data were not publicly available from the authors. The corresponding author may be contacted for questions about data availability.

## Ethics Statement

This study was reviewed and approved by University of Pretoria Research Ethics Committee, Animal Ethics Committee (REC 049-21), Department of Agriculture, Forestry and Fisheries (DAFF) in South Africa (Ref 12/11/1/1/6 (2382SR)) in South Africa, South African National Parks (SANParks), South Africa (Ref: BMTA 006/22).

## Author Contributions

SO, AH and HH conceived the ideas of the study. SO, HH, and AH designed the study. SO, and AH collected the data. SO, AH, KEL and HH designed the methodology. SO, SMM, TM and KEL analysed the data. SO wrote the first draft of the manuscript. All authors contributed significantly to manuscript revision, read, and gave approval for publication.

## Funding

This work was supported by NSF Grant DEB-2106221 through the NSF-NIH-USDA Ecology and Evolution of Infectious Diseases program to W.C.T, P.L.K and H.V.H.

## Acknowledgments

We wish to express our appreciation to the staff of the Skukuza State Veterinary Services for their critical and invaluable support during the experimental component of the research. We also extend our appreciation to the SANParks and all the rangers without whom the passive surveillance system would not work. We also would like to thank Dr Silke Klee of the Robert Koch Institute for the *Bacillus cereus* biovar *anthracis* DNA used as a control in this study. We also like to appreciate Dean J. Herbig for his assistance in the laboratory during this study. Any use of trade, firm, or product names is for descriptive purposes only and does not imply endorsement by the U.S. Government.

## Supplementary Information

Any use of trade, firm, or product names is for descriptive purposes only and does not imply endorsement by the U.S. Government.

**Table S1:** Wildlife species in Kruger National Park, South Africa that tested positive for *Bacillus anthracis* protective antigen (BAPA), lethal factor (*lef*), chromosomal marker (Ba-1) and the capsule region (*capB*) or a combination of these genetic markers and count of animals positive.

| Common name of species | Scientific name of species | Number examined | Percent and number (n) of smears positive for *B. anthracis* by one or more markers |
| --- | --- | --- | --- |
| African buffalo | *Syncerus caffer* | 288 | 38.2%(110) |
| African Elephant | *Loxodonta africana* | 142 | 36.0%(51) |
| African wild dog | *Lycaon pictus* | 1 | 0%(0) |
| Banana bat | *Musonycteris harrisoni* | 1 | 0%(0) |
| Black-backed jackal | *Lupulella mesomelas* | 2 | 0%(0) |
| Blue wildebeest | *Connochaetes taurinus* | 20 | 20.0%(4) |
| Bushbuck | *Tragelaphus scriptus* | 6 | 50.0%(3) |
| Brown hyena | *Hyaena brunnea* | 1 | 0%(0) |
| Chacma Baboon | *Papio ursinus* | 11 | 81.9%(9) |
| Cheetah | *Acinonyx jubatus* | 2 | 0%(0) |
| Common duiker | *Sylvicapra grimmia* | 1 | 0%(0) |
| Common eland | *Taurotragus oryx* | 4 | 25.0%(1) |
| Giraffe | *Giraffa camelopardalis* | 32 | 25.0%(8) |
| Hippopotamus | *Hippopotamus amphibius* | 49 | 34.7%(17) |
| Impala | *Aepyceros melampus* | 525 | 70.3%(369) |
| Greater kudu | *Tragelaphus strepsiceros* | 120 | 76.7%(92) |
| Large-spotted genet | *Genetta tigrina* | 1 | 100%(1) |
| Leopard | *Panthera pardus* | 2 | 0%(0) |
| Lion | *Panthera leo* | 2 | 0%(0) |
| Nile crocodile | *Crocodylus niloticus* | 1 | 100%(1) |
| Nyala | *Tragelaphus angasii* | 47 | 44.7%(21) |
| Ostrich | *Struthio camelus* | 3 | 0%(0) |
| Roan antelope | *Hippotragus equinus* | 29 | 72.5%(21) |
| Spotted hyena | *Crocuta crocuta* | 16 | 0%(0) |
| Steenbok | *Raphicerus campestris* | 11 | 27.3%(3) |
| Common tsessebe | *Damaliscus lunatus* | 4 | 25.0%(1) |
| Vervet monkey | *Chlorocebus pygerythrus* | 4 | 50.0%(2) |
| Warthog | *Phacochoerus africanus* | 10 | 40.0%(4) |
| Waterbuck | *Kobus ellipsiprymnus* | 22 | 41.0%(9) |
| White rhinoceros | *Ceratotherium simum* | 114 | 18.5%(21) |
| Not labelled |  | 150 | 64.0%(96) |
|  | **Total** | **1708** | **890** |

**Table S2:** Positive results of scraped blood smears using *Bacillus anthracis* protective antigen (BAPA), lethal factor (*lef*), chromosomal marker (Ba-1) and the capsule region (*capB*) molecular markers and marker combinations in probe-based real-time/quantitative polymerase chain reaction (qPCR), with “only” indicating exclusive positivity for the respective marker or combination.

| Molecular Markers | Count (%) |
| --- | --- |
| Ba-1 only | 13 (1.5) |
| *lef* only | 165 (18.5) |
| BAPA only | 112(12.6) |
| *capB* only | 5 (0.6) |
| Ba-1+*lef* only | 13(1.5) |
| Ba-1+*capB* only | 3(0.3) |
| Ba-1 + BAPA only | 45(5.1) |
| BAPA + *lef* only | 131(14.7) |
| BAPA + *capB* only | 10(1.1) |
| Ba-1+BAPA+*capB*+*lef* | 393 (44.2) |

## Notes

### Competing Interest Statement

The authors have declared no competing interest.

